# The last frontier for global Non-Communicable Disease action: the Emergency Department – a cross-sectional study from East Africa

**DOI:** 10.1101/2020.07.29.20164632

**Authors:** Christine Ngaruiya, Mbatha Wambua, Thomas Kedera Mutua, Daniel Rafiki Owambo, Morgan Muchemi, Kipkoech Rop, Kaitlin R. Maciejewski, Rebecca Leff, Mūgane Mūtua, Benjamin Wachira

## Abstract

**Background:** Deaths due to non-communicable diseases (NCDs) have surpassed those due to communicable diseases globally and are projected to do so in Africa by 2030. Despite demonstrated effectiveness in high-income country (HIC) settings, the ED is a primary source of NCD care that has been under-prioritized in Africa. In this study, we assess the burden of leading NCDs and NCD risk factors in Kenyan Casualty Department patients, to inform interventions targeting patients with NCDs in emergency care settings.

**Methods:** Using the WHO STEPwise approach to surveillance (STEPS) tool and the Personal Health Questionnaire (PHQ-9), we conducted a survey of 923 adults aged 18 and over at Kenyatta National Hospital Emergency Department (KNH ED), the largest hospital in East Africa between May-October 2018. We used descriptive statistics and covariate-adjusted logistic analysis to analyze results. We included the following socio-demographic variables in our models: age, income, household size (t-test), sex, education, marital status, work status, and poverty status (chi-squared test or fisher’s exact test).

**Findings:** More than a third of respondents had hypertension (35.8%, n=225/628), one in five had raised blood sugar or diabetes (18.3%, n=61/333), and more than one in ten reported having cardiovascular disease (11.7%, n=90/769). Having lower levels of education was associated with tobacco use (OR 6.0, 95% CI 2.808– 12.618, p < 0.0001), while those with higher levels of education reported increased alcohol use (OR 0.620 (95% CI 0.386– 0.994, p = 0. 0472). While a predominant proportion of respondents had had some form of screening for either hypertension (80.3%, n=630/772), blood sugar (42.6%, n=334/767) or cholesterol (13.9%, n=109/766), the proportion of those on treatment was low, with the highest proportion being half of those diagnosed with hypertension reporting taking medication(51.6%, n=116/225). Determinants of disease burden were age, sex, and income.

**Interpretation:** Comprehension of the unique epidemiology and characteristics of patients presenting to the ED is key to guide care in African populations. Patient-driven interventions, and collaboration with community-based stakeholders such as patient navigators, are ideal considerations to sustainably address NCDs leveraging the ED in the resource-limited setting.

**Funding:** Hecht-Albert Global Health Pilot Innovation Award for Junior Faculty, Global Health Leadership Institute, Yale University.

## Introduction

Non-Communicable Diseases (NCDs) annually constitute more than 70% of deaths worldwide.^1^ Furthermore, current disease trends suggest worsening of the situation over the next decade, with the WHO projecting 55 million deaths from NCDs annually by 2030.^2^ NCDs have surpassed communicable diseases as the lead cause of death in all continents except Africa, where NCD-related deaths are nevertheless projected to surpass deaths from communicable diseases, maternal and perinatal conditions, and nutritional deficiencies by 2030.^1^ Eighty-percent of deaths from NCDs occur in low- and middle-income countries (LMICs) with the majority of these occurring prematurely.^2^

The 2013-2020 WHO global action plan for NCDs highlights such targets as: reduction in premature mortality secondary to cardiovascular disease, cancer, diabetes, and chronic respiratory disease; reduction in harmful use of alcohol; reduction in prevalence of tobacco use; reduction in prevalence of raised blood pressure, and increased prevalence of eligible people on appropriate therapy for cardiovascular disease prevention.^2^ Interventions in high-income countries (HICs) have demonstrated the effectiveness of the ED in addressing all of these targets, including tobacco cessation, alcohol cessation, and use of navigators to improve compliance among diabetics, among others.^3-5^ The ED is an optimal setting for these and other novel interventions targeting NCDs.

The ED is also the primary site for presentation of patients with acute NCD-related complications (such as acute coronary syndrome, strokes, diabetic ketoacidosis or asthma exacerbations), mental illness and injury-related complaints.^6^ All of these conditions require timely and effective management in order to mitigate long-term effects of disease. The importance of studying NCDs in the ED and developing high yield interventions to improve management of those presenting with NCD-related acute illness to prevent downstream effects, is paramount.

In this study, we assess the burden of leading NCDs and NCD risk factors in Kenyan Casualty Department patients, in order to inform development of hospital and clinical policies, educational interventions for practitioners on management of NCDs in the emergency setting, and novel interventions targeting patients with NCDs in the Casualty setting. This is the largest epidemiological study on NCDs in an African Emergency Department that we are aware of.

**Research in context**

*Evidence before this study*

Despite demonstrated effectiveness in HIC settings, the ED is a primary source of NCD care that has been under-prioritized in Africa. Although Non-Communicable Diseases (NCDs) have been recognized as major contributors to morbidity and mortality in low- and middle-income countries (LMICs), few estimates of the incidence and epidemiology of leading NCDs and NCD risk factors exist for acute and emergency care settings in Africa. Not only is the ED the site where NCD-related emergencies primarily present, such as acute coronary syndrome or stroke, but also represents a critical setting for novel interventions targeting NCDs.

*Added value of this study*

This is the largest epidemiological study on NCDs in an African Emergency Department that we are aware of. We assessed the burden of leading NCDs and NCD risk factors in Kenyan Casualty Department patients, providing new and robust evidence of the burden of NCDs in EDs in Africa. Our approach using two internationally validated tools, The WHO Stepwise Approach to Surveillance (STEPS) tool and the Personal Health Questionnaire (PHQ-9), is likely generalizable to other low-resource acute and emergency care settings. Our findings demonstrate a high prevalence of NCDs and modifiable NCD risk factors in the Kenyan ED population coupled with a low proportion of patients taking appropriate medications despite diagnosis. A high percentage of our study population reported alcohol or tobacco use, which may be amenable to brief interventions in the ED setting. Several trends demand particular attention. Of these, a double burden of higher prevalence and reduced treatment of NCDs in younger populations in our data, support the need for greater attention to younger adults. We also highlight a disparity in care-seeking behaviors of men, which may not occur unless symptoms present. Such findings demand increased attention for screening and diagnosis of men during clinical encounters, including in the safety net of the ED while continuing to advance community-based prevention efforts. This novel data is crucial for evidence-based development of hospital and clinical policies, educational interventions for practitioners on management of NCDs in the emergency setting, and novel interventions targeting patients with NCDs in acute and emergency care settings.

*Implications of all the available evidence*

This study establishes the ED as a high-risk population with potential for high impact in East Africa, should targeted interventions be implemented. The extensive burden of NCDs and NCD risk factors identified in this population, accompanied by a poor prevalence of patients reporting appropriate medication use, demands further effort to improve the risk reduction, diagnostic processes, and management of NCDs in emergency and acute care settings.

## Methods

### Study Design

The WHO Stepwise Approach to Surveillance (STEPS) tool was used to assess burden of disease of hypertension and diabetes, as well as prevalence of NCD risk factors, and the Personal Health Questionnaire (PHQ-9) was used to assess prevalence of depression.^7,8^ These are both internationally validated tools. Results from the PHQ-9 tool are presented elsewhere.

Two data collectors were hired to assist with administration of surveys. Local data collectors were used who were familiar with the patient population and spoke the native languages. The survey was verbally administered to account for potential barriers with illiteracy, with responses indicated on electronic tablets. Surveys were loaded on to the Kobo software platform for ease of use. The surveys were offered in English and in the national language, Kiswahili. This study received approval from the Institutional Review Board at Yale University.

### Sample size and population

An estimate for the total number of patients seen at the KNH ED in a 3-month period (2014-2015) is between 14,956 and 23, 951, according to Myers et al.^9^ No additional estimate is available to date from the facility or in the literature. Based on this, we had a target sample size of 2,400 or 10% of the upper estimate of total number of presentations over that time period, in line with standard pilot proportions.^10^ We aimed to recruit these 2,400 participants similarly during a 3-month period. However, as a result of system changes during the inception of our study, a large proportion of patients accessing care at the KNH ED were referred away significantly reducing the patient volume. In lieu of our original study plan, we then extended data collection to the entirety of the funding period, for a total of six months and included all patients meeting our inclusion criteria. All patients aged 18 years old or older were considered, which mirrored the age group used in the 2015 Kenya STEPS study.^11^ Patients refusing participation or informed consent were excluded.

### Measures

The analytical sample includes all those who consented to be surveyed. Not all participants answered all questions. Demographic information including age and sex (male, female) were collected. Marital status was categorized as married (currently married or cohabitating), formerly married (divorced, separated, widowed), never married. Participants were asked their highest education level, which was then categorized as secondary school or above (Secondary school completed, High school completed, College/University completed, Post graduate degree), primary school completed, less than primary school completed (less than primary school, no formal schooling). To collect employment information, participants were asked “which of the following best describes your main work status over the past 12 months?” Responses were then categorized as: Employed (Government employee, Non-government employee), Not employed (Homemaker, retired, non-paid, Unemployed (able to work), Unemployed (unable to work), Student, Self-employed.

To gain information about income, participants were asked “Taking the past year, can you tell me what the average earnings of the household have been?” They were also asked “How many people older than 18 years, including yourself, live in your household?”. These variables were then used to create an indicator for household poverty, using the WHO standard. ^12^ The household was considered to be in poverty if household income/household size < 69397.5 KSH (given a poverty cut-off of $1.90 per person per day ($1.9 * 365.25days * 100KShs/$ = 69397.5 KShs). Medication use was determined by asking “are you currently taking [medication]” for a given health disorder. The denominator was restricted only to those who were told they had the related condition by a doctor or other health worker.

### Statistical Analysis

First, descriptive statistics were generated to characterize the overall population. Mean, standard deviation, median, range are presented for numerical variables, percentages are presented for categorical variables, and number missing are presented for all. Bivariate analyses were next conducted for each outcome of interest. Variables were compared by age, income, household size (t-test), sex, education, marital status, work status, and poverty status (chi-squared test or fisher’s exact test). Finally, multivariate analyses were conducted using covariate-adjusted logistic analysis. P < 0.05 was considered statistically significant. Analysis was conducted using SAS 9.4 (SAS Institute, Inc., Cary, NC).

### Role of the funding source

The funders of the study had no role in study design, data collection, data analysis, data interpretation, or writing of the report. The corresponding author had full access to all the data in the study and had final responsibility for the decision to submit for publication.

## Results

There were a total of 923 total respondents, of which 784 (84.9%) provided consent. The mean age was 35y (+/- 13.0y), with a range from 18y to 88y. The majority of respondents were male (61.2%, n= 480/784) (see Table 1). More than half of respondents had completed high school (secondary school) or higher (59.4%, n=466/784). The predominance of respondents reported being married (54.6%, n=428/784). Majority reported being self-employed (41.2%, n=323/784), and 19.9% reported being unemployed (n=156/784). The average reported annual household income was Kenya Shillings (Kshs) 237,888.6, or approximately 2,379 USD, with 30.6% falling below the World Bank international poverty line of 1.90 USD per day (n=240/784).

**Table 1:**
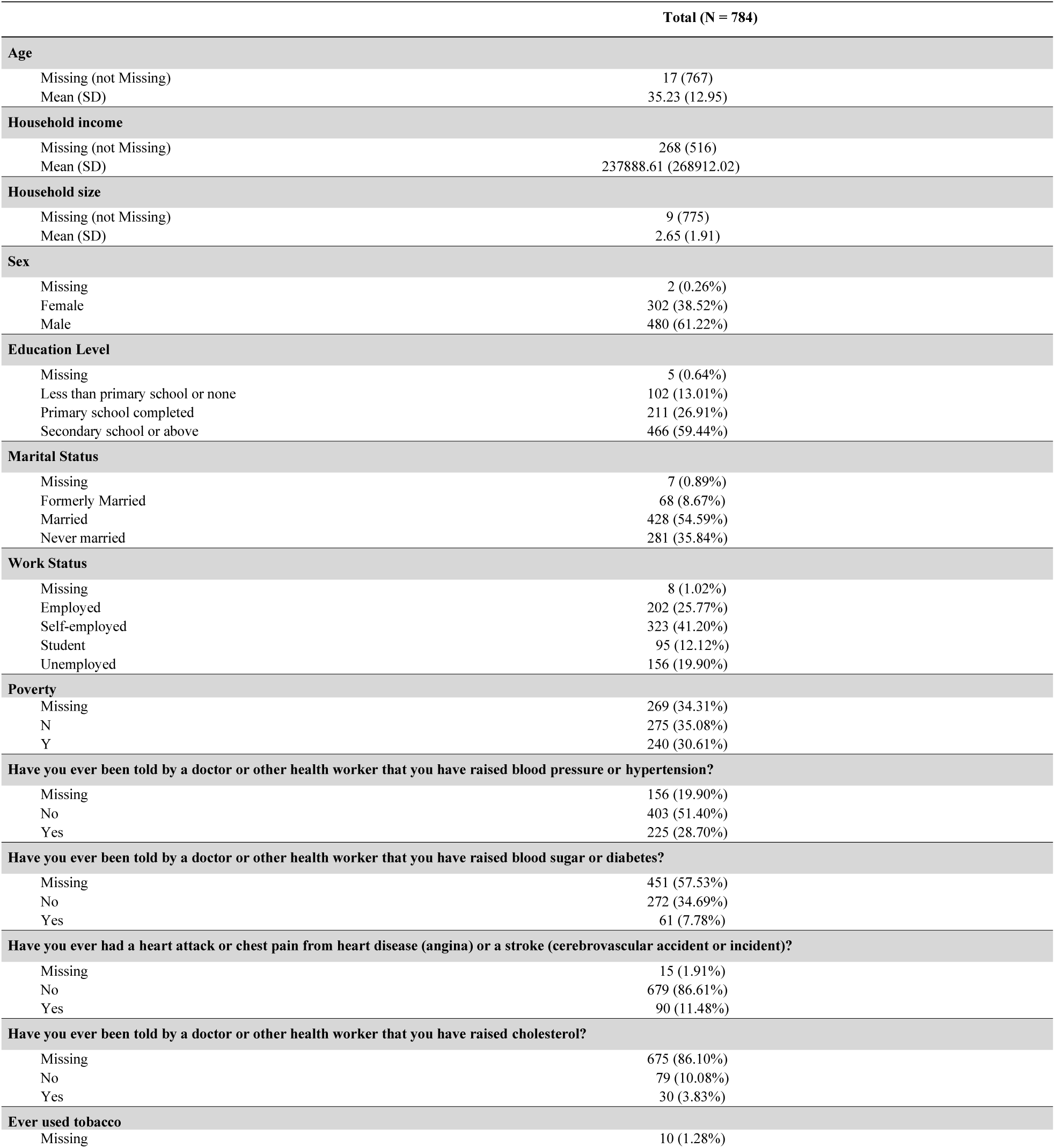
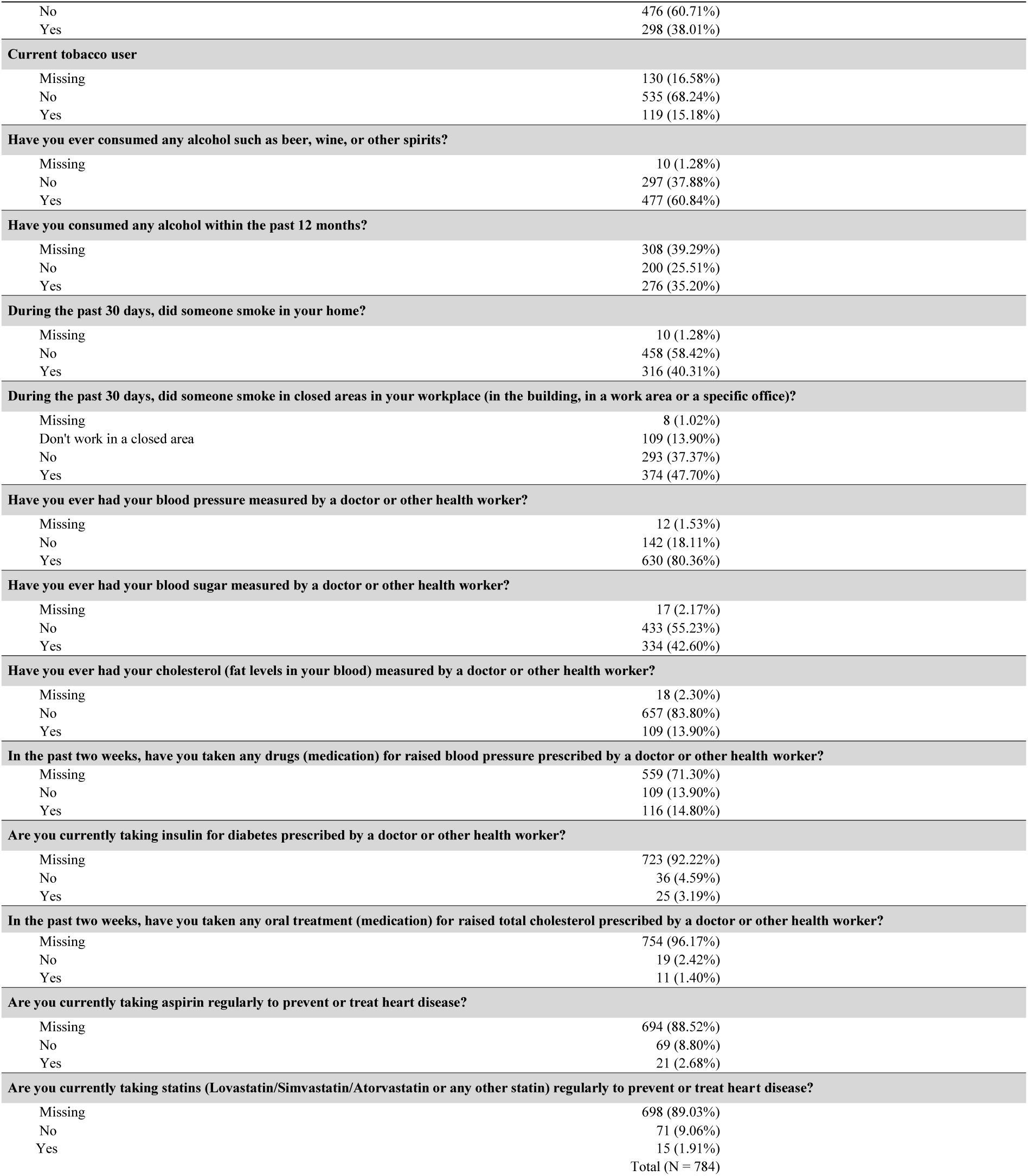
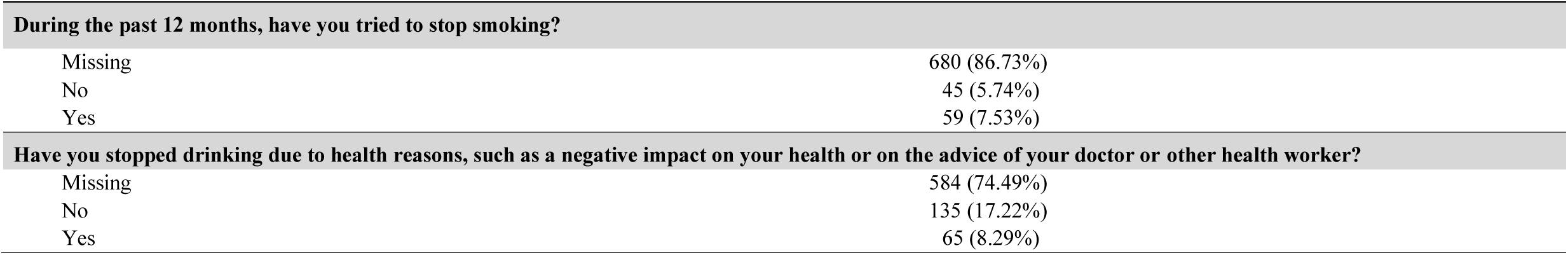
Summary table on participants included in the study.

| Total (N = 784) |  |
| --- | --- |
| Age |  |
| Missing (not Missing) | 17 (767) |
| Mean (SD) | 35.23 (12.95) |
| Household income |  |
| Missing (not Missing) | 268 (516) |
| Mean (SD) | 237888.61 (268912.02) |
| Household size |  |
| Missing (not Missing) | 9 (775) |
| Mean (SD) | 2.65 (1.91) |
| Sex |  |
| Missing | 2 (0.26%) |
| Female | 302 (38.52%) |
| Male | 480 (61.22%) |
| Education Level |  |
| Missing | 5 (0.64%) |
| Less than primary school or none | 102 (13.01%) |
| Primary school completed | 211 (26.91%) |
| Secondary school or above | 466 (59.44%) |
| Marital Status |  |
| Missing | 7 (0.89%) |
| Formerly Married | 68 (8.67%) |
| Married | 428 (54.59%) |
| Never married | 281 (35.84%) |
| Work Status |  |
| Missing | 8 (1.02%) |
| Employed | 202 (25.77%) |
| Self-employed | 323 (41.20%) |
| Student | 95 (12.12%) |
| Unemployed | 156 (19.90%) |
| Poverty |  |
| Missing | 269 (34.31%) |
| N | 275 (35.08%) |
| Y | 240 (30.61%) |
| Have you ever been told by a doctor or other health worker that you have raised blood pressure or hypertension? |  |
| Missing | 156 (19.90%) |
| No | 403 (51.40%) |
| Yes | 225 (28.70%) |
| Have you ever been told by a doctor or other health worker that you have raised blood sugar or diabetes? |  |
| Missing | 451 (57.53%) |
| No | 272 (34.69%) |
| Yes | 61 (7.78%) |
| Have you ever had a heart attack or chest pain from heart disease (angina) or a stroke (cerebrovascular accident or incident)? |  |
| Missing | 15 (1.91%) |
| No | 679 (86.61%) |
| Yes | 90 (11.48%) |
| Have you ever been told by a doctor or other health worker that you have raised cholesterol? |  |
| Missing | 675 (86.10%) |
| No | 79 (10.08%) |
| Yes | 30 (3.83%) |
| Ever used tobacco |  |
| Missing | 10 (1.28%) |
|  | **Total (N = 784)** |
| No | 476 (60.71%) |
| Yes | 298 (38.01%) |
| **Current tobacco user** |  |
| Missing | 130 (16.58%) |
| No | 535 (68.24%) |
| Yes | 119 (15.18%) |
| **Have you ever consumed any alcohol such as beer, wine, or other spirits?** |  |
| Missing | 10 (1.28%) |
| No | 297 (37.88%) |
| Yes | 477 (60.84%) |
| **Have you consumed any alcohol within the past 12 months?** |  |
| Missing | 308 (39.29%) |
| No | 200 (25.51%) |
| Yes | 276 (35.20%) |
| **During the past 30 days, did someone smoke in your home?** |  |
| Missing | 10 (1.28%) |
| No | 458 (58.42%) |
| Yes | 316 (40.31%) |
| **During the past 30 days, did someone smoke in closed areas in your workplace (in the building, in a work area or a specific office)?** |  |
| Missing | 8 (1.02%) |
| Don't work in a closed area | 109 (13.90%) |
| No | 293 (37.37%) |
| Yes | 374 (47.70%) |
| **Have you ever had your blood pressure measured by a doctor or other health worker?** |  |
| Missing | 12 (1.53%) |
| No | 142 (18.11%) |
| Yes | 630 (80.36%) |
| **Have you ever had your blood sugar measured by a doctor or other health worker?** |  |
| Missing | 17 (2.17%) |
| No | 433 (55.23%) |
| Yes | 334 (42.60%) |
| **Have you ever had your cholesterol (fat levels in your blood) measured by a doctor or other health worker?** |  |
| Missing | 18 (2.30%) |
| No | 657 (83.80%) |
| Yes | 109 (13.90%) |
| **In the past two weeks, have you taken any drugs (medication) for raised blood pressure prescribed by a doctor or other health worker?** |  |
| Missing | 559 (71.30%) |
| No | 109 (13.90%) |
| Yes | 116 (14.80%) |
| **Are you currently taking insulin for diabetes prescribed by a doctor or other health worker?** |  |
| Missing | 723 (92.22%) |
| No | 36 (4.59%) |
| Yes | 25 (3.19%) |
| **In the past two weeks, have you taken any oral treatment (medication) for raised total cholesterol prescribed by a doctor or other health worker?** |  |
| Missing | 754 (96.17%) |
| No | 19 (2.42%) |
| Yes | 11 (1.40%) |
| **Are you currently taking aspirin regularly to prevent or treat heart disease?** |  |
| Missing | 694 (88.52%) |
| No | 69 (8.80%) |
| Yes | 21 (2.68%) |
| **Are you currently taking statins (Lovastatin/Simvastatin/Atorvastatin or any other statin) regularly to prevent or treat heart disease?** |  |
| Missing | 698 (89.03%) |
| No | 71 (9.06%) |
| Yes | 15 (1.91%) |
|  | Total (N = 784) |

| During the past 12 months, have you tried to stop smoking? |  |
| --- | --- |
| Missing | 680 (86.73%) |
| No | 45 (5.74%) |
| Yes | 59 (7.53%) |
| Have you stopped drinking due to health reasons, such as a negative impact on your health or on the advice of your doctor or other health worker? |  |
| Missing | 584 (74.49%) |
| No | 135 (17.22%) |
| Yes | 65 (8.29%) |

More than a third of respondents reported being told they had elevated blood pressure or hypertension by a health worker (35.8%, n=225/628) (see Table 2). The average age of those diagnosed was 41.7y (+/- 14.5y), with a female predominance (56.25%, n=126/225) The average reported income among those diagnosed was 2,406.85 USD, and most had completed high school (53.33%, n=120/225) (see Table 1 and 2). In the multivariate analysis, only age, sex, and being below the poverty line were predictors of likelihood of having been diagnosed with hypertension (Table 3). For every 1-year increase in age, the odds of having been told by a doctor or other health worker that they had hypertension increased by 0.066 (6.6%) (95% CI 1.041 – 1.092, p <0.0001). Women had more than double the odds of being told they had hypertension as compared to men (OR 2.335, 95% CI 1.470 – 3.707, p = 0.0003). Those who were below the poverty line had 0.607 (95% CI 0.372– 0.992, p = 0.0462) times the odds of having been told by a doctor or other health worker that they had hypertension, compared to those who were not.

**Table 2:** Sociodemographic characteristics of respondents told by a doctor or other health worker that they had raised blood pressure or hypertension.

|  | No<br>(N = 403) | Yes<br>(N = 225) | Total<br>(N = 628) | P Value |
| --- | --- | --- | --- | --- |
| Age |  |  |  |  |
| Mean (SD) | 33.57 (11.82) | 41.69 (14.48) | 36.51 (13.41) | <0.001*** |
| Household income (Ksh) |  |  |  |  |
| Mean (SD) | 255320.72 (298334.54) | 240685.90 (276783.29) | 249711.30 (290005.41) | 0.62 |
| Household size |  |  |  |  |
| Mean (SD) | 2.60 (2.03) | 2.96 (2.04) | 2.73 (2.04) | 0.034* |
| Sex |  |  |  |  |
| Female | 149 (37.06%) | 126 (56.25%) | 275 (43.93%) | <0.001*** |
| Male | 253 (62.94%) | 98 (43.75%) | 351 (56.07%) |  |
| Education Level |  |  |  |  |
| Less than primary school or none | 34 (8.48%) | 45 (20.00%) | 79 (12.62%) | <0.001*** |
| Primary school completed | 107 (26.68%) | 60 (26.67%) | 167 (26.68%) |  |
| Secondary school or above | 260 (64.84%) | 120 (53.33%) | 380 (60.70%) |  |
| Marital Status |  |  |  |  |
| Formerly Married | 24 (6.02%) | 31 (13.78%) | 55 (8.81%) | <0.001*** |
| Married | 219 (54.89%) | 140 (62.22%) | 359 (57.53%) |  |
| Never married | 156 (39.10%) | 54 (24.00%) | 210 (33.65%) |  |
| Work Status |  |  |  |  |
| Employed | 107 (26.68%) | 53 (23.77%) | 160 (25.64%) | 0.038* |
| Self-employed | 157 (39.15%) | 100 (44.84%) | 257 (41.19%) |  |
| Student | 56 (13.97%) | 16 (7.17%) | 72 (11.54%) |  |
| Unemployed | 81 (20.20%) | 54 (24.22%) | 135 (21.63%) |  |
| Poverty |  |  |  |  |
| No | 135 (54.00%) | 79 (50.64%) | 214 (52.71%) | 0.51 |
| Yes | 115 (46.00%) | 77 (49.36%) | 192 (47.29%) |  |

**Table 3:** Logistic regression analysis on respondents who reported being told that they had hypertension.

| Have you ever been told by a doctor or other health worker that you had hypertension? Yes vs No |  |  |  |  |
| --- | --- | --- | --- | --- |
| Effect |  | Odds Ratio Estimates | 95% Wald Confidence Limits | P-value |
| Sex: | Female vs Male | 2.335 | (1.470, 3.707) | 0.0003 |
| Education Level: | Less than primary school or no formal schooling vs Secondary school or above | 1.109 | (0.492, 2.500) | 0.80 |
| Education Level: | Primary school completed vs Secondary school or above | 1.029 | (0.599, 1.767) | 0.92 |
| Marital Status: | Formerly married vs Never married | 1.813 | (0.702, 4.678) | 0.22 |
| Marital Status: | Married vs Never married | 1.175 | (0.657, 2.102) | 0.59 |
| Work Status: | Employed vs Unemployed | 0.743 | (0.350, 1.576) | 0.44 |
| Work Status: | Self-employed vs Unemployed | 0.630 | (0.313, 1.268) | 0.20 |
| Work Status: | Student vs Unemployed | 0.931 | (0.281, 3.077) | 0.91 |
| Age | Years | 1.066 | (1.041, 1.092) | <.0001 |
| Poverty: | Yes vs No | 0.607 | (0.372, 0.992) | 0.046 |

Nearly one in five patients (18.3%, n=61/333) reported being told they had elevated blood sugar or diabetes by a health worker (see Table 4). The average age among those reporting diagnosis was 49.5y (+/- 13.9y). The mean reported income was 1,851.19 USD. The majority of respondents who reported having been diagnosed with diabetes were men (60.7%, n=37/61), and tended towards lower levels of education with more than half (63.9%, n=39/61) reporting primary school or less. In the multivariate analysis, there was statistically significant evidence of likelihood of diagnosis being associated with advanced age, and for every 1-year increase in age, the odds of having been told that they had raised blood sugar or diabetes increased by 0.072 (7.2%) (95% CI 1.032– 1.113, p =0.0003).

**Table 4:** Sociodemographic characteristics of respondents told by a doctor or other health worker that they had raised blood sugar or diabetes.

|  | No<br>(N = 272) | Yes<br>(N = 61) | Total<br>(N = 333) | P Value |
| --- | --- | --- | --- | --- |
| Age |  |  |  |  |
| Mean (SD) | 38.38 (13.42) | 49.48 (13.89) | 40.43 (14.16) | <0.001*** |
| Household income (Ksh) |  |  |  |  |
| Mean (SD) | 265718.23 (285027.30) | 185119.05 (211180.68) | 250538.12 (274052.37) | 0.041* |
| Household size |  |  |  |  |
| Mean (SD) | 2.62 (1.50) | 3.44 (2.77) | 2.77 (1.83) | 0.027* |
| Sex |  |  |  |  |
| Female | 125 (45.96%) | 24 (39.34%) | 149 (44.74%) | 0.35 |
| Male | 147 (54.04%) | 37 (60.66%) | 184 (55.26%) |  |
| Education Level |  |  |  |  |
| Less than primary school or none | 31 (11.48%) | 20 (32.79%) | 51 (15.41%) | <0.001*** |
| Primary school completed | 75 (27.78%) | 19 (31.15%) | 94 (28.40%) |  |
| Secondary school or above | 164 (60.74%) | 22 (36.07%) | 186 (56.19%) |  |
| Marital Status |  |  |  |  |
| Formerly Married | 30 (11.15%) | 12 (19.67%) | 42 (12.73%) | 0.07 |
| Married | 171 (63.57%) | 40 (65.57%) | 211 (63.94%) |  |
| Never married | 68 (25.28%) | 9 (14.75%) | 77 (23.33%) |  |
| Work Status |  |  |  |  |
| Employed | 73 (27.04%) | 11 (18.33%) | 84 (25.45%) | 0.033* |
| Self-employed | 125 (46.30%) | 33 (55.00%) | 158 (47.88%) |  |
| Student | 22 (8.15%) | 0 (0.00%) | 22 (6.67%) |  |
| Unemployed | 50 (18.52%) | 16 (26.67%) | 66 (20.00%) |  |
| Poverty |  |  |  |  |
| No | 102 (56.67%) | 17 (40.48%) | 119 (53.60%) | 0.06 |
| Yes | 78 (43.33%) | 25 (59.52%) | 103 (46.40%) |  |

38.5% (298/774) of respondents reported having ever used tobacco (smoked or smokeless), and 18.2% (n=119/654) of respondents reported being current smokers (see Table 1). The average age of users was 38y with a range from 17y to 88y, most had completed high school, and the vast majority of users were male (see Table 6). Male sex was a strong predictor of tobacco use, with an eight times (OR 8.86, 95% CI 5.2-15.1) higher odds of use as compared to their female counterparts (see Table 7). Additionally, lower levels of education were more predictive of tobacco use, with those having completed less than primary school being more likely to engage in tobacco use as compared to those that had completed secondary school or above (OR 6.0, 95% CI 2.808– 12.618, p < 0.0001), adjusting for other covariates. 40.8% (n=316/774) of respondents reported being exposed to smoke in home, and 47.7% reported exposure (n=374/776) at work.

**Table 5:** Logistic regression analysis on respondents who reported being told that they had raised blood sugar or diabetes.

| Have you ever been told by a doctor or other health worker that you have raised blood sugar or diabetes? Yes vs No |  |  |  |  |
| --- | --- | --- | --- | --- |
| Effect |  | Odds Ratio Estimates | 95% Wald Confidence Limits | P-value |
| Sex: | Female vs Male | 0.593 | (0.262, 1.346) | 0.21 |
| Education Level: | Less than primary school or no formal schooling vs Secondary school or above | 2.672 | (0.922, 7.744) | 0.082 |
| Education Level: | Primary school completed vs Secondary school or above | 1.251 | (0.481, 3.254) | 0.56 |
| Marital Status: | Formerly married vs Never married | 0.921 | (0.226, 3.756) | 0.94 |
| Marital Status: | Married vs Never married | 0.783 | (0.258, 2.372) | 0.64 |
| Age: | Years | 1.072 | (1.032, 1.113) | 0.0003 |
| Poverty: | Yes vs No | 0.947 | (0.404, 2.221) | 0.90 |

**Table 6:**
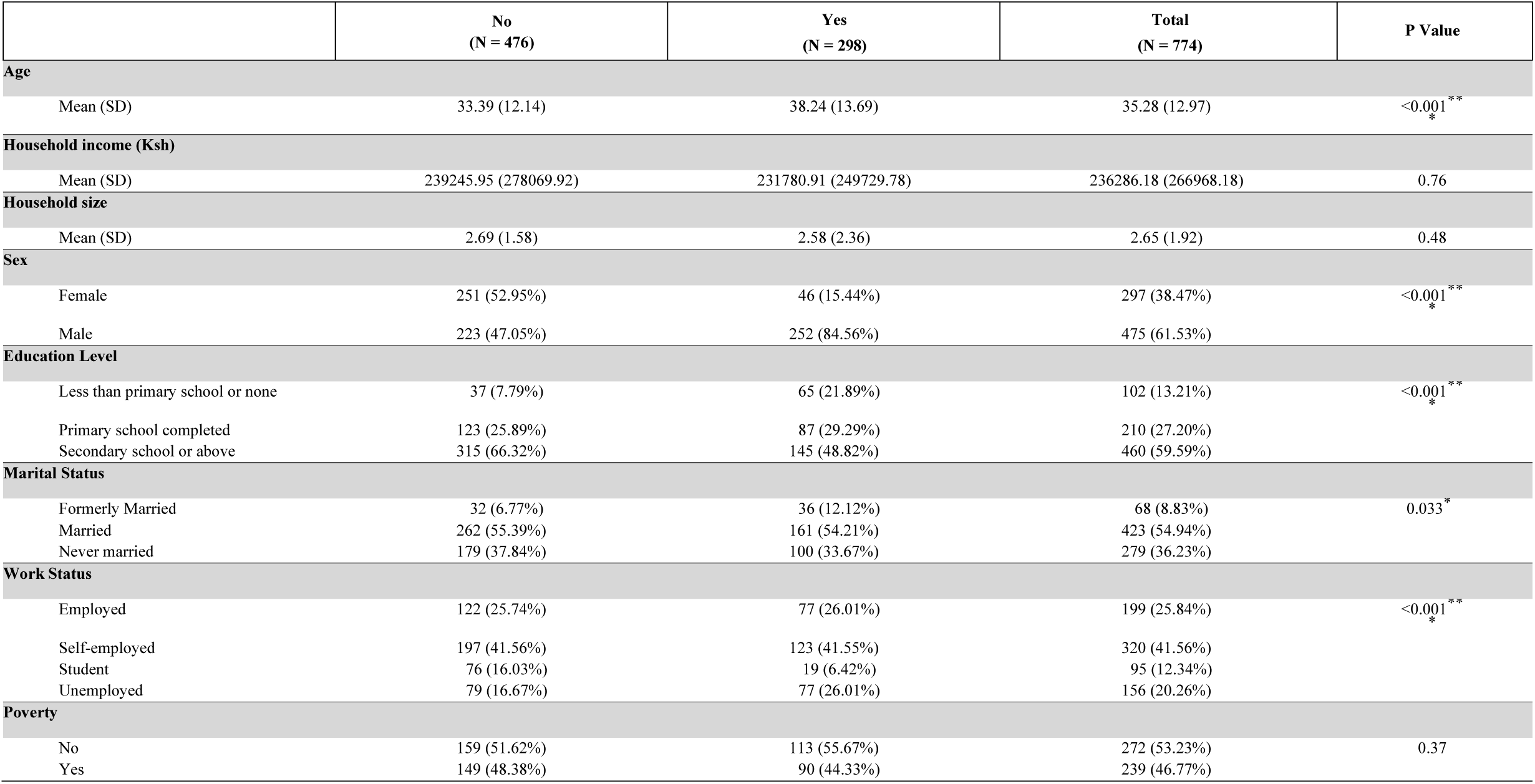
Sociodemographic characteristics of respondents who reported having used tobacco (smoked or smokeless)

|  | No<br>(N = 476) | Yes<br>(N = 298) | Total<br>(N = 774) | P Value |
| --- | --- | --- | --- | --- |
| **Age** |  |  |  |  |
| Mean (SD) | 33.39 (12.14) | 38.24 (13.69) | 35.28 (12.97) | <0.001<sup>**</sup><sub>*</sub> |
| **Household income (Ksh)** |  |  |  |  |
| Mean (SD) | 239245.95 (278069.92) | 231780.91 (249729.78) | 236286.18 (266968.18) | 0.76 |
| **Household size** |  |  |  |  |
| Mean (SD) | 2.69 (1.58) | 2.58 (2.36) | 2.65 (1.92) | 0.48 |
| **Sex** |  |  |  |  |
| Female | 251 (52.95%) | 46 (15.44%) | 297 (38.47%) | <0.001<sup>**</sup><sub>*</sub> |
| Male | 223 (47.05%) | 252 (84.56%) | 475 (61.53%) |  |
| **Education Level** |  |  |  |  |
| Less than primary school or none | 37 (7.79%) | 65 (21.89%) | 102 (13.21%) | <0.001<sup>**</sup><sub>*</sub> |
| Primary school completed | 123 (25.89%) | 87 (29.29%) | 210 (27.20%) |  |
| Secondary school or above | 315 (66.32%) | 145 (48.82%) | 460 (59.59%) |  |
| **Marital Status** |  |  |  |  |
| Formerly Married | 32 (6.77%) | 36 (12.12%) | 68 (8.83%) | 0.033<sup>*</sup> |
| Married | 262 (55.39%) | 161 (54.21%) | 423 (54.94%) |  |
| Never married | 179 (37.84%) | 100 (33.67%) | 279 (36.23%) |  |
| **Work Status** |  |  |  |  |
| Employed | 122 (25.74%) | 77 (26.01%) | 199 (25.84%) | <0.001<sup>**</sup><sub>*</sub> |
| Self-employed | 197 (41.56%) | 123 (41.55%) | 320 (41.56%) |  |
| Student | 76 (16.03%) | 19 (6.42%) | 95 (12.34%) |  |
| Unemployed | 79 (16.67%) | 77 (26.01%) | 156 (20.26%) |  |
| **Poverty** |  |  |  |  |
| No | 159 (51.62%) | 113 (55.67%) | 272 (53.23%) | 0.37 |
| Yes | 149 (48.38%) | 90 (44.33%) | 239 (46.77%) |  |

**Table 7:** Logistic regression analysis on respondents who reported having used tobacco.

| Have you ever used tobacco? Yes vs No |  |  |  |  |
| --- | --- | --- | --- | --- |
| Effect |  | Odds Ratio Estimates | 95% Wald Confidence Limits | P-value |
| Sex: | Female vs Male | 0.117 | (0.069, 0.199) | <.0001 |
| Education Level: | Less than primary school or no formal schooling vs Secondary school or above | 5.952 | (2.808, 12.618) | <.0001 |
| Education Level: | Primary school completed vs Secondary school or above | 1.472 | (0.9, 2.407) | 0.12 |
| Marital Status: | Formerly married vs Never married | 1.083 | (0.435, 2.697) | 0.86 |
| Marital Status: | Married vs Never married | 1.059 | (0.625, 1.795) | 0.83 |
| Work Status: | Employed vs Unemployed | 0.573 | (0.272, 1.209) | 0.14 |
| Work Status: | Self-employed vs Unemployed | 0.594 | (0.295, 1.197) | 0.15 |
| Work Status: | Student vs Unemployed | 0.362 | (0.127, 1.028) | 0.056 |
| Age | Years | 1.020 | (0.999, 1.042) | 0.059 |
| Poverty: | Yes vs No | 0.664 | (0.423, 1.04) | 0.074 |

Another 61.6% (477/774) reported having used alcohol, majority being current users, having consumed alcohol within the past year. The average age of alcohol users was 36y, and majority of users were also male (see Table 8). Females have 0.324 (95% CI 0.215– 0.488, p <.0001) times the odds of ever consuming any alcohol, compared to males, adjusting for other covariates (see Table 9). Those who completed primary school have 0.620 (95% CI 0.386– 0.994, p = 0. 0472) times the odds, compared to those who attended secondary school or above.

**Table 8:**
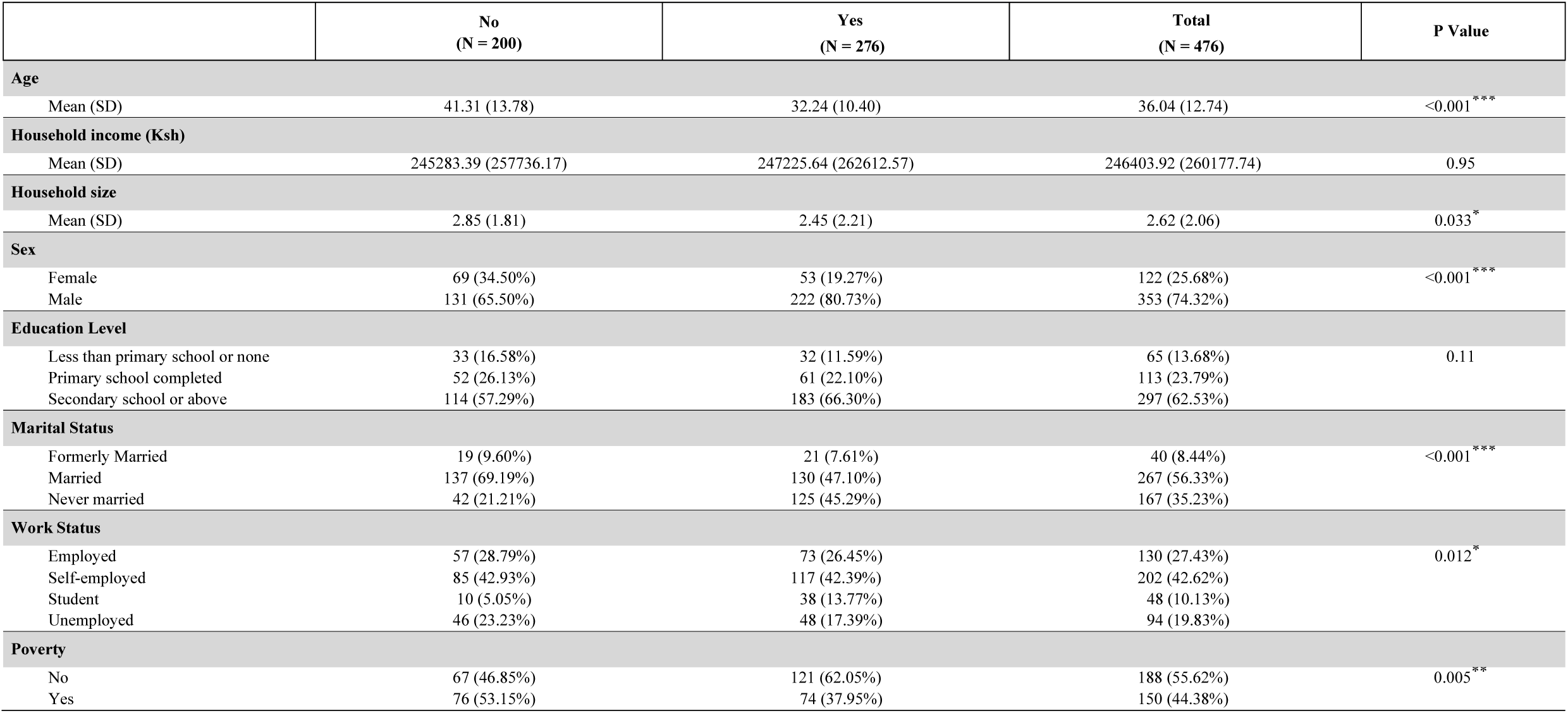
Sociodemographic characteristics of respondents who reported having consumed any alcohol within the past 12 months.

|  | No<br>(N = 200) | Yes<br>(N = 276) | Total<br>(N = 476) | P Value |
| --- | --- | --- | --- | --- |
| Age |  |  |  |  |
| Mean (SD) | 41.31 (13.78) | 32.24 (10.40) | 36.04 (12.74) | <0.001*** |
| Household income (Ksh) |  |  |  |  |
| Mean (SD) | 245283.39 (257736.17) | 247225.64 (262612.57) | 246403.92 (260177.74) | 0.95 |
| Household size |  |  |  |  |
| Mean (SD) | 2.85 (1.81) | 2.45 (2.21) | 2.62 (2.06) | 0.033* |
| Sex |  |  |  |  |
| Female | 69 (34.50%) | 53 (19.27%) | 122 (25.68%) | <0.001*** |
| Male | 131 (65.50%) | 222 (80.73%) | 353 (74.32%) |  |
| Education Level |  |  |  |  |
| Less than primary school or none | 33 (16.58%) | 32 (11.59%) | 65 (13.68%) | 0.11 |
| Primary school completed | 52 (26.13%) | 61 (22.10%) | 113 (23.79%) |  |
| Secondary school or above | 114 (57.29%) | 183 (66.30%) | 297 (62.53%) |  |
| Marital Status |  |  |  |  |
| Formerly Married | 19 (9.60%) | 21 (7.61%) | 40 (8.44%) | <0.001*** |
| Married | 137 (69.19%) | 130 (47.10%) | 267 (56.33%) |  |
| Never married | 42 (21.21%) | 125 (45.29%) | 167 (35.23%) |  |
| Work Status |  |  |  |  |
| Employed | 57 (28.79%) | 73 (26.45%) | 130 (27.43%) | 0.012* |
| Self-employed | 85 (42.93%) | 117 (42.39%) | 202 (42.62%) |  |
| Student | 10 (5.05%) | 38 (13.77%) | 48 (10.13%) |  |
| Unemployed | 46 (23.23%) | 48 (17.39%) | 94 (19.83%) |  |
| Poverty |  |  |  |  |
| No | 67 (46.85%) | 121 (62.05%) | 188 (55.62%) | 0.005** |
| Yes | 76 (53.15%) | 74 (37.95%) | 150 (44.38%) |  |

**Table 9:**
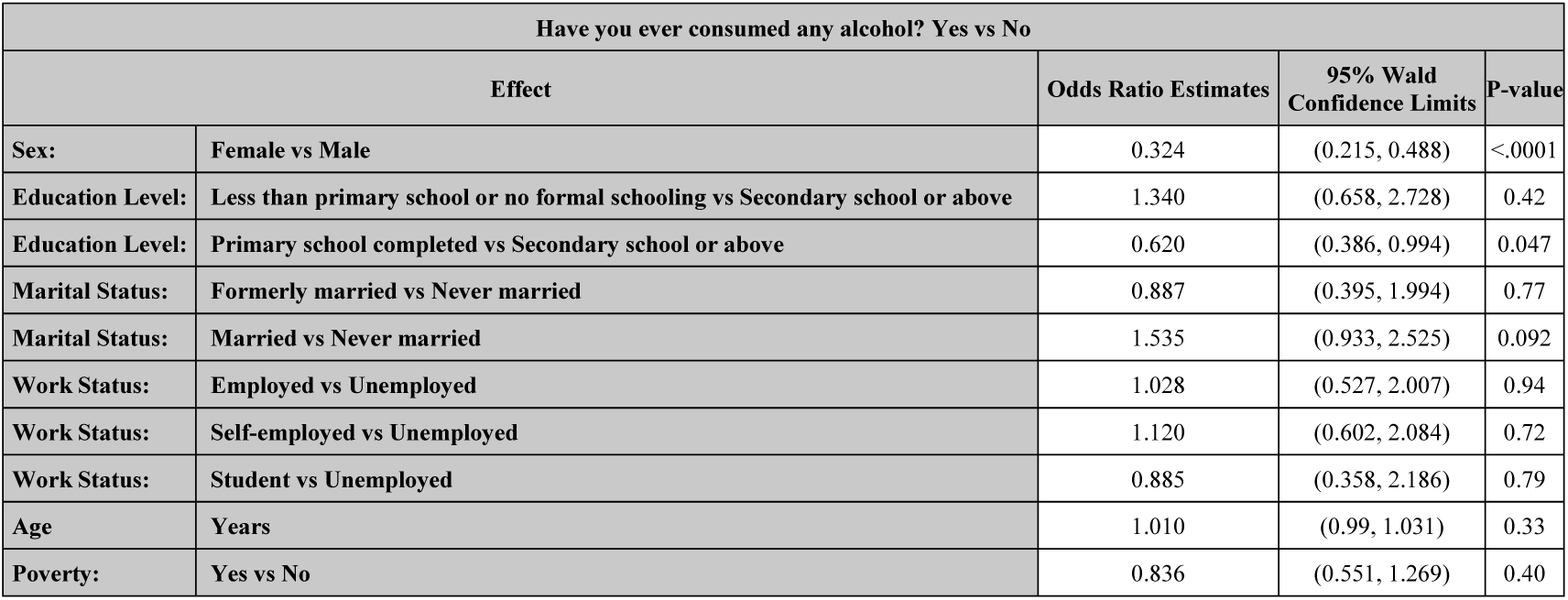
Logistic regression analysis on respondents who reported having ever consumed alcohol.

Majority of respondents had either had their blood pressure (80.3%, n=630/772), blood sugar (42.6%, n=334/767) or cholesterol (13.9%, n=109/766) measured (see Tables 10-12). Of those that had been told they had elevated blood pressure, 51.6% (n=116/225) reported taking medications. Age was the only sociodemographic variable that had statistically significant evidence predicting likelihood of using antihypertensives (see Table 13). For every one year increase in age, the odds of ever having taken medication for hypertension in the past two weeks increased by 0.089 (8.9%) (95% CI 1.044– 1.136, p <0.0001), adjusting for other covariates. Furthermore, of those that had been told they had elevated blood sugar or diabetes, 41% (n=25/61) reported taking insulin. Again, age was the only predictor with statistical evidence for association with taking insulin in diabetic patients, with the odds of use increasing with age (OR 1.3, 95%CI 1.1 - 1.6, p = 0.0115), adjusting for other covariates (see Table 14). There were no statistically evident predictors of taking statins or aspirin. While more than one in ten respondents reported a history of cardiovascular disease – either angina, heart disease or stroke (11.7%, n=90/769), only one in four (23.3%, n=21/90) of these were taking aspirin and one in six (17.44%, n=15/86) were taking a statin. Only 3 CVD patients were taking both an aspirin and statin.

**Table 10:** Sociodemographic characteristics of respondents who reported having ever had their blood pressure measured by a doctor or other health worker.

|  | No<br>(N = 142) | Yes<br>(N = 630) | Total<br>(N = 772) | P Value |
| --- | --- | --- | --- | --- |
| Age |  |  |  |  |
| Mean (SD) | 29.70 (8.68) | 36.53 (13.43) | 35.25 (12.95) | <0.001*** |
| Household income (Ksh) |  |  |  |  |
| Mean (SD) | 195892.40 (164019.98) | 248629.58 (289709.91) | 238102.79 (270034.14) | 0.016* |
| Household size |  |  |  |  |
| Mean (SD) | 2.28 (1.22) | 2.73 (2.04) | 2.65 (1.92) | <0.001*** |
| Sex |  |  |  |  |
| Female | 24 (16.90%) | 275 (43.79%) | 299 (38.83%) | <0.001*** |
| Male | 118 (83.10%) | 353 (56.21%) | 471 (61.17%) |  |
| Education Level |  |  |  |  |
| Less than primary school or none | 22 (15.49%) | 79 (12.58%) | 101 (13.12%) | 0.55 |
| Primary school completed | 40 (28.17%) | 168 (26.75%) | 208 (27.01%) |  |
| Secondary school or above | 80 (56.34%) | 381 (60.67%) | 461 (59.87%) |  |
| Marital Status |  |  |  |  |
| Formerly Married | 10 (7.04%) | 56 (8.95%) | 66 (8.59%) | 0.009** |
| Married | 65 (45.77%) | 360 (57.51%) | 425 (55.34%) |  |
| Never married | 67 (47.18%) | 210 (33.55%) | 277 (36.07%) |  |
| Work Status |  |  |  |  |
| Employed | 39 (27.46%) | 160 (25.56%) | 199 (25.91%) | 0.14 |
| Self-employed | 60 (42.25%) | 259 (41.37%) | 319 (41.54%) |  |
| Student | 23 (16.20%) | 72 (11.50%) | 95 (12.37%) |  |
| Unemployed | 20 (14.08%) | 135 (21.57%) | 155 (20.18%) |  |
| Poverty |  |  |  |  |
| No | 57 (55.88%) | 214 (52.45%) | 271 (53.14%) | 0.53 |
| Yes | 45 (44.12%) | 194 (47.55%) | 239 (46.86%) |  |

**Table 11:** Sociodemographic characteristics of respondents who reported having ever had their blood sugar measured by a doctor or other health worker.

|  | No<br>(N = 433) | Yes<br>(N = 334) | Total<br>(N = 767) | P Value |
| --- | --- | --- | --- | --- |
| Age |  |  |  |  |
| Mean (SD) | 31.19 (10.11) | 40.43 (14.14) | 35.18 (12.85) | <0.001*** |
| Household income (Ksh) |  |  |  |  |
| Mean (SD) | 225626.40 (257991.74) | 250538.12 (274052.37) | 236562.06 (265185.17) | 0.29 |
| Household size |  |  |  |  |
| Mean (SD) | 2.55 (1.99) | 2.77 (1.82) | 2.64 (1.92) | 0.11 |
| Sex |  |  |  |  |
| Female | 148 (34.34%) | 150 (44.91%) | 298 (38.95%) | 0.003** |
| Male | 283 (65.66%) | 184 (55.09%) | 467 (61.05%) |  |
| Education Level |  |  |  |  |
| Less than primary school or none | 50 (11.55%) | 51 (15.36%) | 101 (13.20%) | 0.15 |
| Primary school completed | 113 (26.10%) | 95 (28.61%) | 208 (27.19%) |  |
| Secondary school or above | 270 (62.36%) | 186 (56.02%) | 456 (59.61%) |  |
| Marital Status |  |  |  |  |
| Formerly Married | 24 (5.56%) | 42 (12.69%) | 66 (8.65%) | <0.001*** |
| Married | 210 (48.61%) | 212 (64.05%) | 422 (55.31%) |  |
| Never married | 198 (45.83%) | 77 (23.26%) | 275 (36.04%) |  |
| Work Status |  |  |  |  |
| Employed | 114 (26.33%) | 84 (25.45%) | 198 (25.95%) | <0.001*** |
| Self-employed | 159 (36.72%) | 158 (47.88%) | 317 (41.55%) |  |
| Student | 72 (16.63%) | 22 (6.67%) | 94 (12.32%) |  |
| Unemployed | 88 (20.32%) | 66 (20.00%) | 154 (20.18%) |  |
| Poverty |  |  |  |  |
| No | 151 (52.98%) | 119 (53.60%) | 270 (53.25%) | 0.89 |
| Yes | 134 (47.02%) | 103 (46.40%) | 237 (46.75%) |  |

**Table 12:** Have you ever had your cholesterol (fat levels in your blood) measured by a doctor or other health worker?

|  | No<br>(N = 657) | Yes<br>(N = 109) | Total<br>(N = 766) | P Value |
| --- | --- | --- | --- | --- |
| Age |  |  |  |  |
| Mean (SD) | 33.95 (12.06) | 42.74 (15.33) | 35.16 (12.91) | <0.001*** |
| Household income (Ksh) |  |  |  |  |
| Mean (SD) | 224326.85 (248126.70) | 324800.00 (364173.96) | 239160.48 (270350.41) | 0.024* |
| Household size |  |  |  |  |
| Mean (SD) | 2.57 (1.62) | 3.17 (3.19) | 2.65 (1.93) | 0.06 |
| Sex |  |  |  |  |
| Female | 247 (37.71%) | 51 (46.79%) | 298 (39.01%) | 0.07 |
| Male | 408 (62.29%) | 58 (53.21%) | 466 (60.99%) |  |
| Education Level |  |  |  |  |
| Less than primary school or none | 88 (13.41%) | 13 (12.04%) | 101 (13.22%) | 0.53 |
| Primary school completed | 180 (27.44%) | 25 (23.15%) | 205 (26.83%) |  |
| Secondary school or above | 388 (59.15%) | 70 (64.81%) | 458 (59.95%) |  |
| Marital Status |  |  |  |  |
| Formerly Married | 51 (7.80%) | 14 (12.96%) | 65 (8.53%) | 0.001** |
| Married | 350 (53.52%) | 71 (65.74%) | 421 (55.25%) |  |
| Never married | 253 (38.69%) | 23 (21.30%) | 276 (36.22%) |  |
| Work Status |  |  |  |  |
| Employed | 163 (24.92%) | 33 (30.56%) | 196 (25.72%) | 0.37 |
| Self-employed | 270 (41.28%) | 46 (42.59%) | 316 (41.47%) |  |
| Student | 86 (13.15%) | 9 (8.33%) | 95 (12.47%) |  |
| Unemployed | 135 (20.64%) | 20 (18.52%) | 155 (20.34%) |  |
| Poverty |  |  |  |  |
| No | 226 (52.31%) | 45 (60.00%) | 271 (53.45%) | 0.22 |
| Yes | 206 (47.69%) | 30 (40.00%) | 236 (46.55%) |  |

**Table 13:** Logistic regression analysis on respondents who reported having taken medication for hypertension.

| In the past two weeks have you taken medication for hypertension? Yes vs No |  |  |  |  |
| --- | --- | --- | --- | --- |
| Effect |  | Odds Ratio Estimates | 95% Wald Confidence Limits | P-value |
| Sex: | Female vs Male | 0.524 | (0.233, 1.181) | 0.12 |
| Education Level: | Less than primary school or no formal schooling vs Secondary school or above | 1.314 | (0.381, 4.538) | 0.67 |
| Education Level: | Primary school completed vs Secondary school or above | 0.889 | (0.353, 2.241) | 0.80 |
| Marital Status: | Formerly married vs Never married | 0.482 | (0.108, 2.145) | 0.34 |
| Marital Status: | Married vs Never married | 0.621 | (0.222, 1.734) | 0.36 |
| Work Status: | Employed vs Unemployed | 0.565 | (0.158, 2.027) | 0.38 |
| Work Status: | Self-employed vs Unemployed | 0.854 | (0.272, 2.680) | 0.79 |
| Work Status: | Student vs Unemployed | 0.332 | (0.024, 4.523) | 0.41 |
| Age: | Years | 1.089 | (1.044, 1.136) | <.0001 |
| Poverty: | Yes vs No | 1.254 | (0.515, 3.051) | 0.62 |

**Table 14:** Logistic regression analysis on respondents who reported taking ins.

| Are you currently taking insulin? Yes vs No |  |  |  |  |
| --- | --- | --- | --- | --- |
| Effect |  | Odds Ratio Estimates | 95% Confidence Limits | p-Value |
| Sex: | Female vs Male | 0.806 | (0.095, 6.862) | 0.84 |
| Education Level: | Less than primary school or no formal schooling vs Secondary school or above | 0.063 | (0.001, 2.779) | 0.15 |
| Education Level: | Primary school completed vs Secondary school or above | 1.433 | (0.07, 29.185) | 0.81 |
| Marital Status: | Formerly married vs Never married | 0.277 | (0.007, 11.35) | 0.50 |
| Marital Status: | Married vs Never married | 0.061 | (0.001, 2.541) | 0.14 |
| Age: | Years | 1.32 | (1.064, 1.637) | 0.012 |
| Poverty: | Yes vs No | 4.187 | (0.274, 63.935) | 0.30 |

## Discussion

In this study, we described the burden of NCDs and mental health at the Kenyatta National Hospital Emergency Department, the largest hospital in East Africa. More than a third of respondents had hypertension, one in five had raised blood sugar or diabetes, and more than one in ten reported having cardiovascular disease. More than one third reported tobacco use, and two thirds reported alcohol use. Majority reported not taking medications despite diagnosis, with the highest proportion being half of those diagnosed with hypertension reporting taking medication. Determinants of disease burden were age, sex, and income. Having lower levels of education was associated with tobacco use, however those with higher levels of education reported alcohol use. While a predominant proportion of respondents had had some form of screening for either hypertension, diabetes, or high cholesterol, the proportion of those on treatment was low. Our results showed a systematically higher NCD burden among the ED population as compared to the national population as demonstrated by the Kenya national Stepwise approach to Surveillance (STEPs) study conducted 2 years prior^11^ where the prevalence of hypertension was only in a quarter of the population, less than 3% had raised blood sugar or diabetes, only 13% reported tobacco use and 19% reported alcohol use. The mental health results from our study will be presented elsewhere.

There was a higher prevalence of hypertension in our study on an ED population (35.8%) as compared to the national STEPs study (24.5%) utilizing the same validated survey tool,^13^ and as compared to the prevalence of national populations in the surrounding region: 24.3% in Uganda,^14^ 26% in Tanzania, ^15^ and 11.2% in Rwanda.^16^ The same is true for diabetes with only 2.1% of the general population reporting a diabetes diagnosis nationally in the Kenya STEPs study.^11^ This high prevalence of disease in the ED population is well-established in the US, Canada and UK settings.^17^ While the population presenting to the ED are likely to have higher prevalence of disease as this is a facility-based setting, and results may be biased by patients seeking care with known disease, this still establishes the ED as a high-risk population with potential for high-impact if targeted interventions are implemented, particularly in populations that may not otherwise access the healthcare system. A systematic review and meta-analysis by Armitage et al demonstrates the opportunity for detection of hypertension during screening in the ED.^17^ This in turn provides the opportunity for patient education, linkage to care, or prescription of medication through the ED, and could affect immediate care such as screening for end-organ damage during the acute visit.^4^

The average age of those diagnosed with hypertension was 42y. This is in contrast to US-based populations where hypertension is predominantly prevalent in those 60 years and older, as demonstrated by data from the National Health and Nutrition Examination Survey (NHANES).^18^ However, this observation of afflicted younger populations is not uncommon in African populations. Results from the Rwanda STEPS demonstrated a hypertension prevalence of 13% among 25-34 year olds, and 19% among 35-44 year olds.^16^ The case is comparable for diabetes, with average age of diagnosis in our sample being 49y. Similarly, in the Rwanda case, the distribution of diabetes did not show significant difference for younger individuals. Our data support the need for greater attention to younger adults in this setting, as opposed to recommendations traditionally targeting only those 40 years and older.^19^ By implementing primary and secondary prevention efforts for these younger groups the costly implications of NCD complications can be mitigated.

Women had a higher likelihood of hypertension diagnosis. The odds of women reporting a higher prevalence of disease is divergent from most African countries where there is a male predominance.^20^ In the national STEPS study, men had much higher likelihood of not having had their blood pressure taken (71.1%, 95% CI 64.9-77.3) as compared to women (41.3%, 95% CI 36.5-46.0). Just as in our ED sample population, there was also a higher proportion of women that reported having been diagnosed as compared to men. Contrary to this, there was a predominance of men who reported having been diagnosed with diabetes (60.7%) in our study, whereas in the national data, prevalence of disease was equal among both sexes. These findings highlight an interesting and important disparity in care-seeking behaviors of men, which may not occur unless symptoms exist. This is particularly detrimental for hypertension that tends to present asymptomatically.^21^ To that end, increased attention is needed for screening and diagnosis of men during clinical encounters, including in the safety net of the ED while continuing to advance community-based prevention efforts.

Those below the poverty line had 40% less likelihood of being diagnosed with hypertension. Additionally, among all patients that reported having a diagnosis of hypertension, diabetes, high cholesterol or CVD, majority had a secondary school education or higher. These markers of socioeconomic status raise concerns about diagnosis and management of disease among marginalized populations in the ED. The *Lancet* Commission on Reframing NCDs and Injuries (NCDI) for the Poorest Billion highlights poverty as a primary driver for NCDs.^22^ In our study, the average reported annual household income was Kenya Shillings (Kshs) 237,888.6, or approximately 2,379 USD, with 30.6% falling below the World Bank international poverty line of 1.90 USD per day (n=240/784). The Lancet Commission has facilitated development of “national NCDI poverty commissions”, as well as leveraging the WHO PEN-Plus package – two interventions occurring at the policy level that could help facilitate sustainable change for individuals affected by NCDs in LMICs.^23^

One in five Kenyans reported having used tobacco (21.3%) in the national STEPS study,^11^ whereas 38.5% reported use in our study. Nearly half of individuals reported exposure to smoke in the workplace or at home. In sum, exposure to tobacco is a significant problem in this population, and an alarming burden of disease is attributable to tobacco alone.^24^ The WHO Tobacco Free Initiative highlights the role for brief tobacco interventions in healthcare settings, which have demonstrated effectiveness in US-based ED populations,^5^ but there are no studies to date on interventions in an African ED.

Overall, the prevalence of alcohol use was remarkably high. Those with higher levels of education were more likely to report engaging in alcohol use. This observation is divergent from tobacco use, which is more likely among those with lower levels of education. This phenomenon of education being associated with increased alcohol use has been demonstrated in other settings, including the US, however the downstream effects of alcohol use such as morbidity and mortality of disease remains disproportionately higher in those of lower socioeconomic status.^25^ and public health programming should be targeted accordingly. Brief interventions on alcohol intake in clinical settings are another intervention with demonstrated success in the ED setting.^3^

Among those diagnosed with hypertension or diabetes, two of the leading NCDs, the vast majority were not taking medications for disease. In our study, the only determinant of using medications for NCDs and NCD risk factors was age. However, the lack of statistical evidence of differences in medication use compared across socioeconomic determinants likely indicates a universal lack of access regardless of sex, education and poverty level. Alternatively, it is possible that differences were not detected in some cases due to relatively small sample sizes for these questions. Our findings highlight the need to develop interventions to increase appropriate use of medication for individuals younger than 50y. Prescription of antihypertensives at ED discharge has been shown to improve blood pressure with no significant detrimental side effects in follow-up.^26^ Mobilizing the ED to initiate, and help ensure compliance with, medications should be prioritized.

### Next steps

Clinical protocols should be developed and routinely reviewed with providers, to help standardize therapy and empower education on self-care practices for patients. Education on hypertension and diabetes can also be facilitated by staff, and task-shifting can be employed such as through use of navigators - dedicated, trained nurses or lay health workers. Additionally, linkage to care programs such as those connecting to Community Health Workers could be implemented.^27^ Mobile health (mHealth) technology presents a unique low-cost, highly effective opportunity to enhance health education, support self-monitoring, and improve follow-up,^28^ which could also be leveraged in ED populations. National policies that further address availability and affordability of medications are needed, in addition to ensuring enforcement of the WHO essential medicines list with first-line blood pressure and diabetes medications. In addition, efforts beyond the ED include media campaigns, and strengthening primary healthcare. WHO initiatives such as the SAFER technical package address alcohol access, and include strengthening restrictions on alcohol availability, enforcing restrictions on alcohol advertising and promotion, among other recommendations.^29^ Finally, our findings highlight the importance of legislation that implements and enforces smoke-free zones in public places and workplaces, as well as regulates how these policies are enforced with clear sanctions for entities not abiding by guidelines.^30^

### Limitations

The sample studied in our population may not have findings generalizable to the national population given the study was conducted at a tertiary, referral hospital which may represent a population with higher prevalence of comorbidities. With that said, in selecting a pilot study site, we felt that this site was likely to provide one of the most optimal sites to capture patients presenting from across the region, and our results provide evidence of concerning disease prevalence in a large population that seeks care here. Furthermore, our study provides insight into some of the most economically challenged populations in Kenya who receive care in the public health system, which is where majority commonly seek care in Kenya and Africa at large.

## Conclusion

The ED acts as a catchment site for patients that may not otherwise frequent a healthcare setting. The unique epidemiology and characteristics of patients presenting to the ED are key in order to guide care. Opportunities exist for further research including longitudinal data collection through surveillance and registries to better understand epidemiology of disease, as well as implementation science to guide effective intervention planning. Patient-driven interventions, and collaboration with community-based stakeholders such as CHWs, would be ideal considerations to sustainably address NCDs leveraging the ED in the resource-limited setting.

## Data Availability

All data used in this study are publicly available and are referenced in the manuscript.

## Contributors

CN and BW initially designed the study. MW and RL refined the study design and developed the conception or design of the work. TK, DO, M Muchemi, KP and M Mutua contributed substantially to the acquisition of data. KM analyzed and interpreted data for the work. CN drafted the manuscript with MW, BW, and RL. All authors interpreted data and critically revised the manuscript. All authors have approved the final version for publication.

## Acknowledgements

The authors would like to acknowledge the contributions to data analysis by Mr. Manuel Martinez, Mr. Jesse Martinez, and also would like to thank the Yale Center for Analytical Sciences (YCAS) support staff.

## Declaration of interests

We declare no competing interests.

## Data Sharing

All data used in this study are publicly available and are referenced in the manuscript.

## Supplemental Tables

## Notes

### Competing Interest Statement

The authors have declared no competing interest.

### Funding Statement

This study received funding from the Hecht-Albert Global Health Pilot Innovation Award for Junior Faculty, Global Health Leadership Institute, Yale University.

### Author Declarations

This study received approval from the Institutional Review Board at Yale University.

